# Localised community circulation of SARS-CoV-2 viruses with an increased accumulation of single nucleotide polymorphisms that adversely affect the sensitivity of real-time reverse transcription assays targeting Nucleocapsid protein

**DOI:** 10.1101/2021.03.22.21254006

**Authors:** Catherine Moore, Louise Davies, Rhiannydd Rees, Laura Gifford, Heather Lewis, Amy Plimmer, Andrew Barratt, Nicole Pacchiarini, Joel Southgate, the COG-UK Consortium, Matthew J. Bull, Joanne Watkins, Sally Corden, Thomas R. Connor

## Abstract

Currently the primary method for confirming acute SARS-CoV-2 infection is through the use of molecular assays that target highly conserved regions within the viral genome. Many, if not most of the diagnostic targets currently in use were produced early in the pandemic, using genomes sequenced and shared in early 2020. As viral diversity increases, mutations may arise in diagnostic target sites that have an impact on the performance of diagnostic tests. Here, we report on a local outbreak of SARS-CoV-2 which had gained an additional mutation at position 28890 of the nucleocapsid protein, on a background of pre-existing mutations at positions 28881, 28882, 28883 in one of the main circulating viral lineages in Wales at that time. The impact of this additional mutation had a statistically significant impact on the Ct value reported for the N gene target designed by the Chinese CDC and used in a number of commercial diagnostic products. Further investigation identified that, in viral genomes sequenced from Wales over the summer of 2020, the N gene had a higher rate of mutations in diagnostic target sites than other targets, with 115 issues identified affecting over 10% of all cases sequenced between February and the end of August 2020. In comparison an issue was identified for ORFab, the next most affected target, in less than 1.4% of cases over the same time period. This work emphasises the potential impact that mutations in diagnostic target sites can have on tracking local outbreaks, as well as demonstrating the value of genomics as a routine tool for identifying and explaining potential diagnostic primer issues as part of a laboratory quality management system. This work also indicates that with increasing genomic sequencing data availability, there is a need to re-evaluate the diagnostic targets that are in use for SARS-CoV-2 testing, to better target regions that are now demonstrated to be of lower variability.

## Introduction

Since the emergence of SARS-CoV-2 in December 2019 and the declaration of a pandemic in March 2020, there has been significant interest in the rate of change across the viral genome and the implication that these changes might have in terms of transmission and pathogenicity.

While SARS-CoV-2 acquires mutations less frequently than Influenza (1,2) it has nonetheless been possible to utilise genomic epidemiology to identify and monitor seeding events and transmission clusters in granular detail, especially once genomic data is integrated with epidemiological information. The opportunity afforded to monitor the emergence of a new virus into a susceptible population has led to an international effort to sequence SARS-CoV-2 samples to support local pandemic response and global public health research (3). In the UK, this effort has been co-ordinated by the COG-UK consortium (4) which as of the 30^th^ July 2020, had sequenced 39519 viruses from across the UK. The Public Health Wales, Pathogen Genomics Unit (PenGU) itself had contributed 6757 sequences as of the 30th of July, representing almost 40% of all confirmed Welsh COVID-19 cases as of that date.

Currently the primary method for confirming acute infection is through the use of molecular assays that target highly conserved regions within the viral genome. The global sharing of the first SARS-CoV-2 genome on the 10th of January enabled the rapid development of diagnostic molecular tests, targeting regions of the virus that were expected to show low levels of variation. Since January 2020, the WHO have produced lists of approved assays that have formed the basis of laboratory defined tests (LDTs) (5). These assays have, in many cases, gone on to form the blueprint of many commercially available assays. Due to the rapid spread of SARS-CoV-2, and the need for large volumes of testing globally, many of the tests in use are derived from the very earliest SARS-CoV-2 genomes, and do not take into account the virus diversity that has accumulated in the first 8 months of the pandemic, which has the potential to introduce mutations that may impact diagnostic tests.

Common targets include the open reading frame (ORFab), envelope protein (E), the nucleocapsid protein (NP) and Spike protein (S). Most commercial assays employ a multiplex approach, targeting two or more genes in combination with an internal control. This offers a highly sensitive and specific approach to virus detection, meaning that if one target fails, then that will not automatically produce a false-negative. Sensitivity of assays however varies considerably (6)(7), particularly at the lower level of detection (LLOD) with fluctuation seen between the different gene targets (8) and different platforms. Currently, there is no WHO international standard available to which assays can be verified against.

One significant risk to assay performance is the emergence of SARS-CoV-2 viruses with mutations that directly affect the target region of any given assay (9),(10). The effects of such mutations depend on where the mutation occurs, with those affecting the 5’ end of a primer having much less effect than those that affect the 3’ end or probe. The availability of very large numbers of publically available SARS-CoV-2 complete genome sequences means that surveillance is in place to monitor for changes that might adversely affect assay sensitivity. Generally however, these platforms report on global incidence of any given change as a proportion of the total present in the dataset, and although regional incidence is reported, localised changes in incidence may be missed. Some systems (such as CoV-GLUE (11)) automatically identify mutations that fall within published primer sets, however, this poses two problems. Firstly, the organisations who should be monitoring these changes may not be aware of CoV-GLUE and similar systems, and this monitoring takes time and effort, which may be problematic in the midst of a pandemic. Secondly, the presence of a mutation in a primer region doesn’t necessarily mean that the mutation will impact the assay -for that, data relating to the Cycle Threshold (Ct) value for each target assessed for each sequenced genome would be required. This data is not routinely available, and so this makes the assessment of genomic changes on diagnostic tests on global sequence data collections impossible to perform, other than by those who collect this data and have access to the relevant sequence for collected samples.

Within Wales, changes that could affect diagnostic assays are continuously monitored, and because the majority of samples sequenced in Wales were initially processed by PHW diagnostic laboratories, the data required to identify and quantify the effect of genome sequence variation on diagnostic assays are readily available. Here, we describe the emergence of a SARS-CoV-2 viral variant in a region of Wales that acquired a mutation which affects the primer region of the Chinese CDC nucleocapsid assay (12), which has a measurable effect on the performance of the associated diagnostic assay. These viruses have emerged from a widespread UK transmission lineage that already possess three mutations within the same primer region, and these results have implications for SARS-CoV-2 assay design, review and validation.

## Methods

During June 2020, a cluster of SARS-CoV-2 cases was investigated in a town in South East Wales. As part of the investigation mass screening was undertaken of 900 individuals.

Dry throat swabs (13) were self-collected under the guidance of a community sampling team and the samples were returned to the Wales Specialist Virology Centre, University Hospital of Wales for processing.

On receipt in the laboratory, swabs were broken into 0.9 mL of guanidinium thiocyanate-based lysis buffer as previously described (13). Total nucleic acid purification was undertaken using the fully automated Perkin Elmer (Waltham, Mass, USA) Chemagic 360 followed by amplification using the Perkin Elmer SARS-CoV-2 RT-PCR following manufacturer’s instructions (12,14). The RT-PCR itself was performed on a ThermoFisher QuantStudio 5 instrument.

All of the positive results are routinely analysed to determine the proportion of samples suitable for referring for whole genome sequencing based on the threshold crossing point (Ct) of each gene target NP and ORF1. Early analysis of SARS-CoV-2 in Wales saw a Ct threshold of 30 set as the maximum permissible Ct for samples to be sequenced. During this analysis it was observed that there was an unusual pattern in the results, with the NP assay reducing in sensitivity by at least 3 cycles when compared to the ORF1 assay and the positive control.

Further investigation was undertaken of previous routine assay runs to determine if this pattern was a common observation. Where it was identified, details on the regional location of the patient who submitted the sample were noted.

As the mass screening had been initiated in response to an increase of cases locally, genomic data was already available for the earlier cases associated with the incident. It was therefore possible to rapidly identify the likely transmission lineage present in the region and interrogate specifically the changes occurring across the NP coding region to determine if the pattern of results observed could be attributed to specific mutations. In total, 51 positive cases were identified that had undergone genome sequencing, and were available for analysis.

Genomic sequencing on all samples as part of this study was performed as per the ARTIC protocol, V3 (15). Diagnostic residual samples were reverse transcribed and amplified as per the ARTIC protocol. The resulting amplicons were then subject to library preparation using Nextera XT, as per the manufacturers’ instructions, followed by sequencing on a NextSeq 500 sequencing instrument using 2×150 cycle kits, multiplexing up to 384 samples per run.

Following sequencing samples were processed using a containerised, nextflow version of the ARTIC bioinformatics pipeline developed for Illumina (15,16). Samples were then uploaded to a dedicated COVID-19 environment on MRC CLIMB (17) where samples are integrated with the wider UK dataset (18) and are classified into SARS-CoV-2 lineages (19) and UK transmission groups using the COG-UK phylogenetics pipeline. UK Transmission groups are defined using ancestral state reconstruction which seeks to identify groups of isolates whose ancestor is inferred to be from the UK, as described previously (20). Because of sample bias in the global sequencing dataset, these clusters may represent more than one introduction into the UK, but provide a useful basis for the subdivision of cases into meaningful groups that can then form the basis of further epidemiological investigation. These results were then pulled back into PHW where they are integrated with other data using data-flo (21) and visualised using Microreact (17,22). Assembled sequences were processed by a CoV-GLUE instance running in the COVID-19 environment on MRC CLIMB to identify SNPs or features that fell within documented primer sites. The report generated by CoV-GLUE was then checked and issues were summarised for the complete dataset.

## Results

Initial examination of the recorded Ct values for assays of cases related to the local incident showed no discernable reduction in Ct value when compared to the other targets in the assay (Figure S1), a shift of between 3-4 Ct values were observed, and in some cases no amplification at all was observed in the NP assay with the other targets not being adversely affected. Following this observation a genomic analysis was undertaken to examine mutations against public primer sets in Welsh data. Interrogation of the genomic data from samples associated with the incident and also from across Wales identified that the SARS-CoV-2 viruses associated with the incident being investigated fell within the global B1.1 lineage. In total, 4373/5326 high quality sequences sequenced in Wales between February and August fall within B.1 and its sublineages. Of the 4373 samples falling into B.1 and its sublineages, 2570 cases from Wales by the end of August were known to possess three SNPs (G28881A, G28882A, G28883C), which are characteristic of a number of UK transmission lineages within this group. These SNPs all fall at the 3’ end of the forward primer targeting the nucleocapsid gene in the assay described by the Chinese CDC. The position of this particular series of 3 SNPs has suggested that the changes would have minimal effect on assay sensitivity. This analysis identified that the cases associated with the local incident additionally all possessed a fourth SNP (C28890T) that falls closer to the middle of the primer (Table ST1), upstream of the three existing SNPs found in the samples from B1 which was likely to be the cause of further primer destabilisation. All of the samples associated with the incident that possessed the extra SNP fell within a single UK transmission group, UK805 (Figure 1).

**Figure 1.**
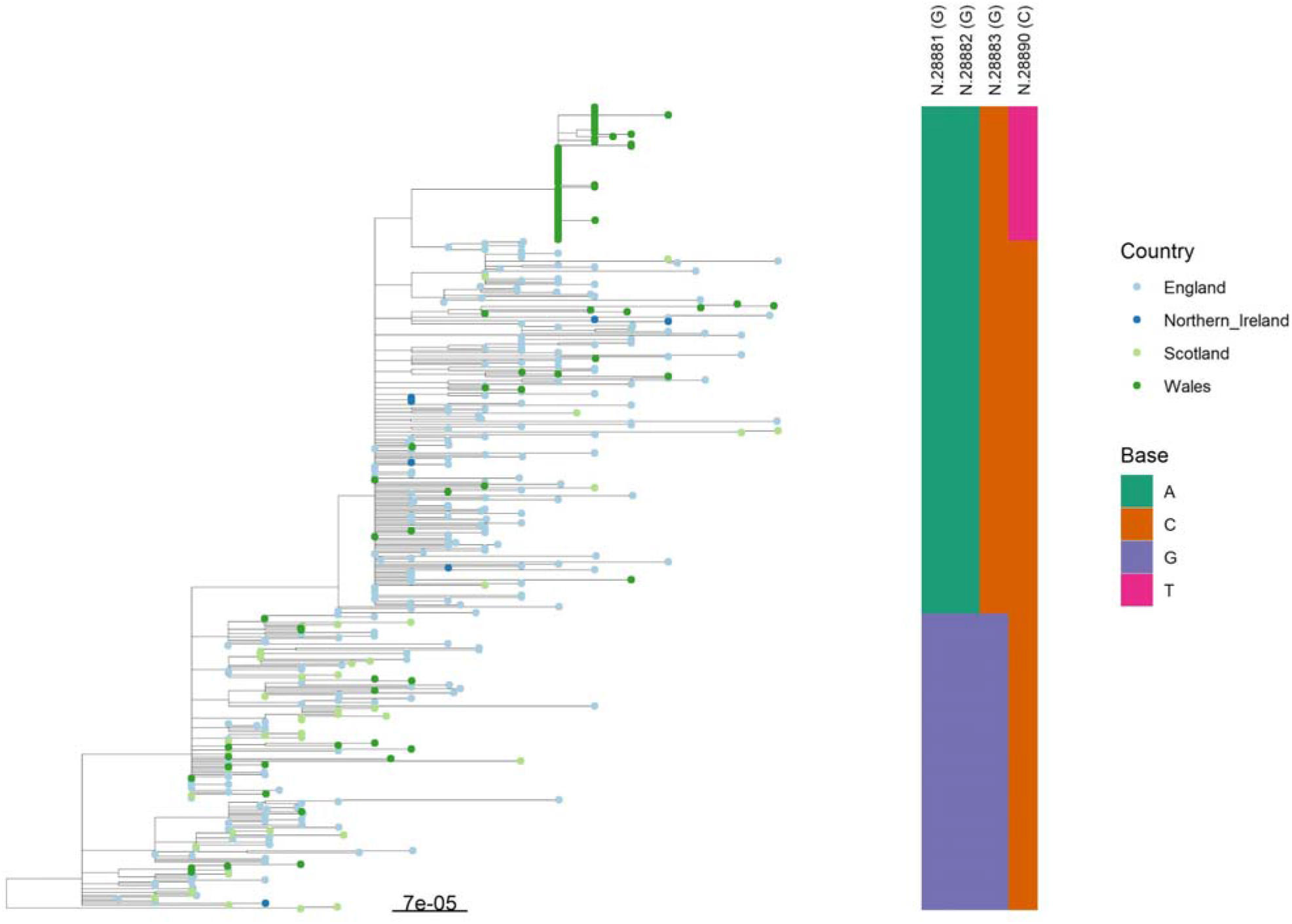
Representative phylogeny of UK SARS-CoV-2 diversity. Included tree tips are a sample of 10% of all tips, weighted by UK lineage abundance, with all UK805 samples included. Tips are coloured by country of sample origin and base at positions 28881, 28882, 28883 and 28890 of MN908947.3 are indicated (vertical bar).

Analysis of the cases on the COG-UK analysis platform revealed that UK805 included both Welsh and English cases, and that UK805 has some substructure. The COG-UK phylogenetics pipeline provides a system for subdividing transmission groups based on tree structure, which within COG-UK is named a phylotype. A phylotype is effectively a numerical representation of tree branching. All samples associated with the incident were classified into a single phylotype, UK805_1.2. This phylotype is exclusively found in Wales, however, a closely related virus, sharing a common ancestor with the Welsh cluster of cases is also found elsewhere in England, and also possesses the fourth mutation (Figure 2). Wider analysis of all whole genome sequences from across Wales confirmed that the fourth mutation is largely localised to one particular region of South East Wales (Figure 3).

**Figure 2.**
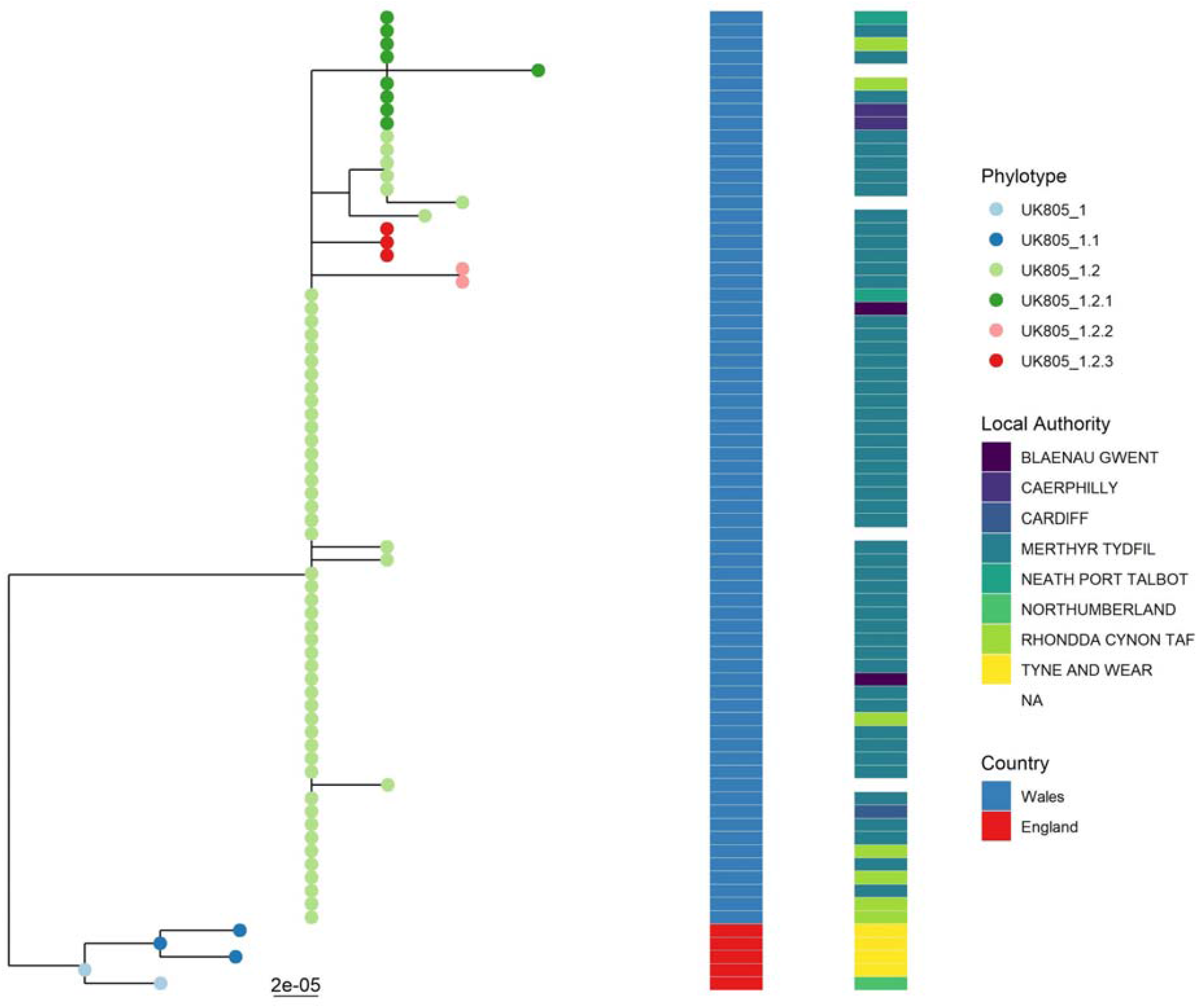
UK805 phylogeny indicating phylotype prefix (tip points), country of sample origin (left hand vertical bar) and resident local authority (right hand vertical bar).

**Figure 3.**
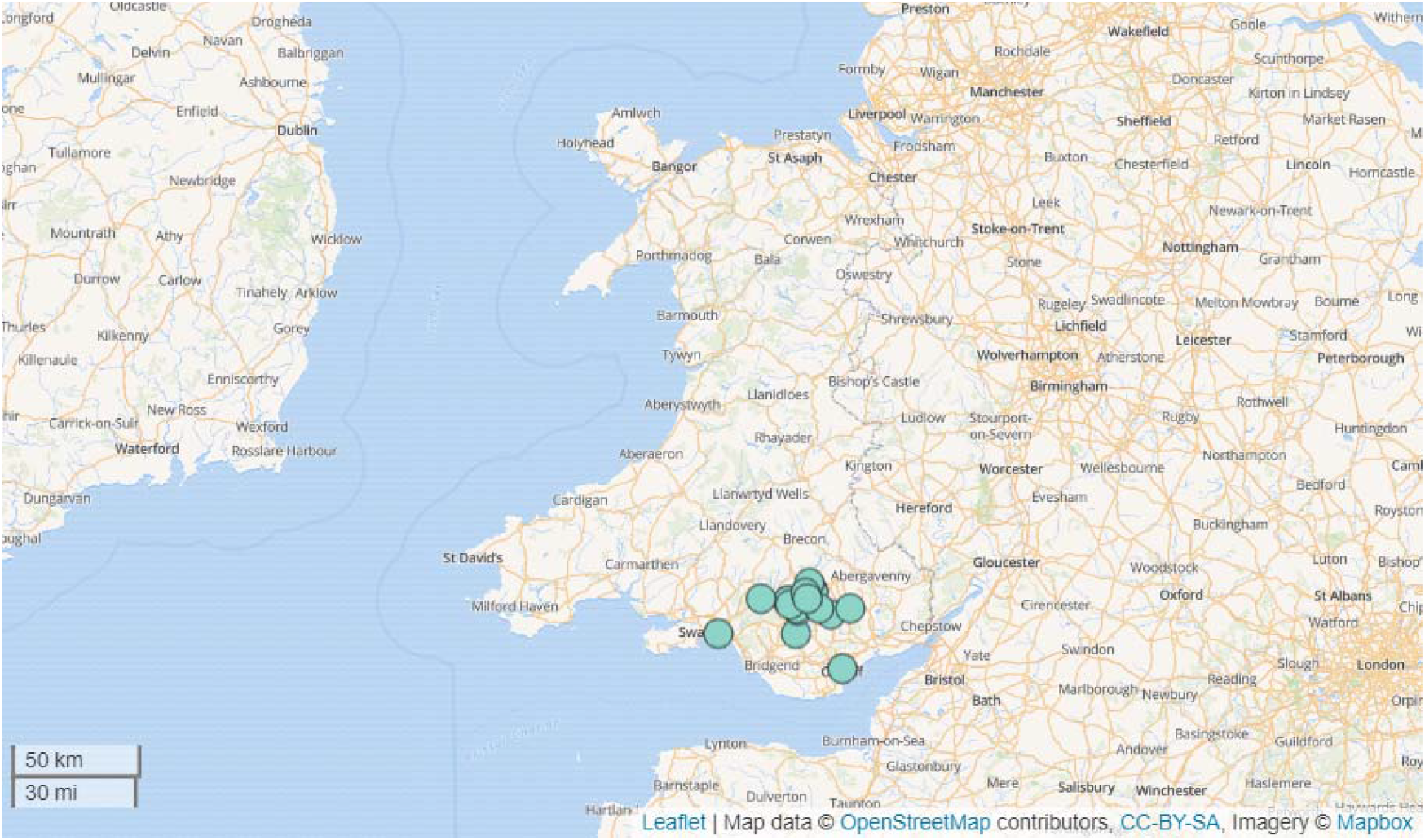
Welsh resident location of cases within lineage UK805.

Of 51 cases from this region recorded between 17th May 2020 and 16th July 2020, 100% had the identified SNP. This particular variant is identified in both community and hospitalised cases, and was first observed in Wales sequence data on the 15th May 2020 (Figure 4).

**Figure 4.**
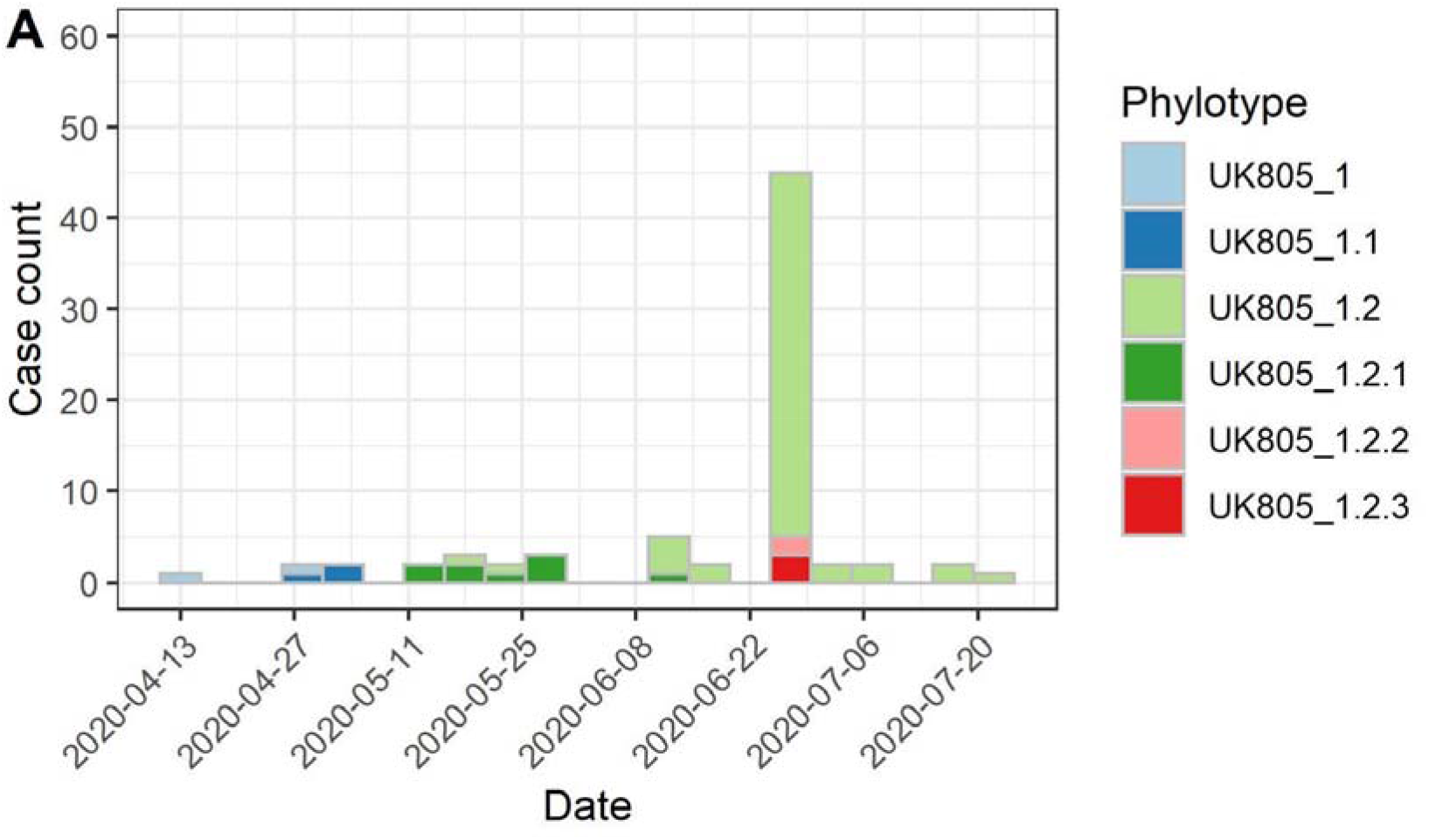
Count of UK805 phylotypes sequenced in England and Wales

Having identified that the incident group covers a single monophyletic group, and all have the fourth SNP, we sought to examine the effect of the extra SNP on the results of the diagnostic tests performed. In total, sequence data and diagnostic Cts from the diagnostic assays using the Chinese CDC primers were available for the 51 cases that formed part of the incident. As a comparator, we identified all cases sequenced between June and the end of August 2020 that had been tested using a diagnostic assay that made use of the Chinese CDC primers, and which had the three SNPs but not the fourth SNP, for which we had results available (n=120, tables with Ct values in Supplementary information). Comparing the Ct values for the N gene target between these groups revealed a difference in the median Ct for the N gene target (24 vs 28 in the incident group with the fourth SNP), which is statistically significant by a Mann Whitney U test (p-value 0.001). The median for the second target used in these diagnostic assays (targeting Orf1ab) was not statistically different between the groups (24 vs 23 in the incident group with the fourth SNP).

A review of all assays used in Wales that referenced in the manufacturer’s instructions the Chinese CDC assay was undertaken to determine whether a similar effect on the sensitivity of the NP gene assay was noted. Where the assay origin wasn’t referenced, samples with this NP gene change were tested through the assay to determine effect. This review showed that assays produced by Luminex (Austin Texas, USA) (both NxTAG assay and the ARIES assay) and the SeeGene (Seoul, S Korea) SARS-CoV-2 (generation 1 and 2) were affected as well as the Perkin Elmer assay already described.

In addition to the issue identified within the specific primers against the nucleocapsid gene, highlighted by the local incident, our analysis reveals 150 mutations/sets of mutations in the primer sites for other assays targeting the nucleocapsid gene developed by the CDC, the Thailand Ministry of Health, the Japanese National Institute of Infectious Diseases and HKU in our sequenced data collected between February and August 2020. The number of issues associated with nucleocapsid primers is considerably higher than those targeting other genes, with the 115 mutations/combinations in diagnostic sites for the N gene comparing to 7 mutations/combination of mutations in diagnostic primers located in the E Gene, 20 in ORFab and 8 in RdRP (Table ST1).

The absolute effect on numbers of cases affected is similarly striking, even excluding the impact of cases from B1.1 possessing the three mutations at the start of the CDC N primer. Excluding samples with the 3 mutations in the N gene primer, there were 730 other sequenced viruses in our dataset with one or more mutations in the N gene, representing 13.7% of all sequenced Welsh cases between February and August 2020. This contrasts with the number of viruses with mutations in diagnostic primer sites elsewhere in the SARS-CoV-2 genome. Examining the entire dataset for all other gene targets, the 730 viruses with mutations in diagnostic primers in the N gene is ten times the number of viruses found with a mutation/mutations in the target with the next most frequent number of issues (ORFab – 73 cases with issues, Figure 5).

**Figure 5.**
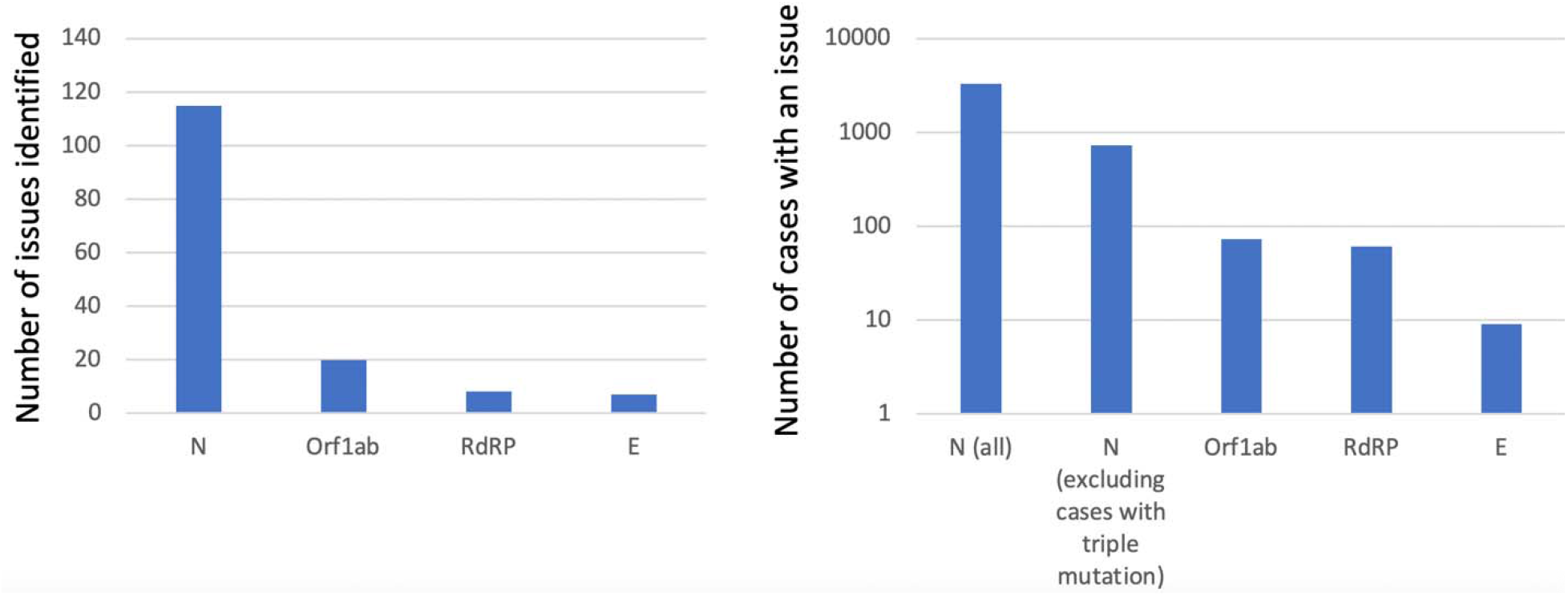
showing the number of sequenced cases with one or more mutations in diagnosti primers by region, and the number of unique mutations/combinations of mutations observed in diagnostic assays in the sequenced Welsh data

## Discussion

Whole genome sequencing is being performed at an unprecedented scale globally to monito the transmission patterns of SARS-CoV-2. This data has also afforded the opportunity to perform rapid assessment of the effect of mutations on the frontline molecular assays used to diagnose acute infection. Although this is notionally the responsibility of manufacturers, in a pandemic situation, normal processes may not be suitable for the monitoring of changes and their effects on diagnostic assays. The generation of large quantities of sequence data, linked to diagnostic Ct values provides an opportunity for laboratories to proactively examine the performance of assays to assure themselves of the continued appropriateness of assays, based on the circulating virus population, and to inform interpretation of test results.

Changes to SARS-CoV-2 have already been reported that may affect various published assays at varying degrees, however, here we report one of the first cases of the circulation of a SARS-CoV-2 virus that possesses mutations that have directly impacted adversely on the sensitivity of NP assays based specifically on the Chinese CDC assay (4).

The significance of this change is most likely to affect post-acute positive samples in assays that target either NP alone or are part of a multiplex with a second gene, such as those produced by Perkin Elmer and Luminex ARIES that both target just NP and ORF1. Indeed, we have already seen positive cases that report only in the ORF1 assay.

The initial change was observed on analysis of the assay results themselves, highlighting the ongoing need to review results before reporting. However, once the change was observed it was the availability of the whole genome sequencing from across Wales to allow rapid assessment of mutations across the NP gene that might account for the change in sensitivity.

Fortunately, the manufacturer affected did reference the most likely assay used as the blueprint for the commercial test. This meant it was possible to be specific about where the mutation had occurred and why the assay was so adversely affected, and then to also review assays in use that also reference the Chinese CDC assay.

However, an increasing number of manufacturers are producing assays that are not based on previously published primer sets and so monitoring for mutations that might affect sensitivity is more challenging. By understanding assay kinetics through robust local verification prior to introduction into routine service, followed by systematic review of all results prior to reporting, this should highlight any unusual results that might suggest mutations affecting the assay.

Data from publically available whole genome sequences primarily offers the commercial company the opportunity to confirm if changes have occurred in the affected region as part of their own surveillance. As we have shown, it also allows for rapid and ongoing assessment of mutations to be identified in viruses as they emerge across the commonly targeted regions of commercial assay, whether or not they reference a specific assay. This type of routine checking of primers should be a core activity for manufacturers, using data that is publicly available, however, there appears to have been no information communicated by the manufacturers to indicate that issues could exist with their assays.

The SARS-CoV-2 virus identified with this pattern of SNPs is currently geographically limited to one region of South East Wales, however there is evidence of low level ongoing transmission in the local community. It is likely therefore, that should there be a significant increase in cases in the coming winter, this virus may become more common across Wales. Furthermore, this additional mutation has occurred on a genetic backbone that is found across a lineage that represents the most frequently sequenced lineage in the UK. More generally, the large number of changes affecting NP across different assays may also point to the fact that NP is not a stable diagnostic target for testing. In the case of viruses from lineage B1.1, should there be further accumulation of SNPs in the NP, then there is a strong chance that any assay based on the Chinese CDC NP primers will become largely redundant. Collectively this work demonstrates the value of genomic data to identify the source of issues with PCR-based diagnostic tests. This work also demonstrates that sequence information can be effectively used as part of laboratory quality management to rapidly identify issues in tests and allow corrective measures to be put in place locally. This work also demonstrates that the publicly available sequence data shared as part of the COVID-19 pandemic response provides ample data to commercial companies to design better probes and identify potential issues before they have a chance to affect patients. Although the responsibility sits with commercial companies to review the effect of mutations on probes, there is little evidence of this occurring. With the sequence data available there is an unprecedented amount of information to support this. While challenging for labs and commercial providers of diagnostic tests, this data provides a powerful tool to ensure that tests are fit for purpose and keep up with changes in the viral genome as the pandemic progresses.

## Supporting information

COG-UK Authorship List

## Data Availability

All sequence data used is provided in the COG-UK bioproject PRJEB37886
Samples used in the analysis are included in supplementary table S1

https://www.ebi.ac.uk/ena/browser/view/PRJEB37886

## Acknowledgements

T.R.C. acknowledges support from the MRC, which funded the computational resources used by the project (grant reference MR/L015080/1), as well as specific funding from Welsh Government, which provided funds for the sequencing and analysis Welsh samples used in this study, via Genomics Partnership Wales. Welsh sequencing has also been supported by the COVID-19 Genomics UK Consortium (COG-UK), as well as using genomics data that has partly been funded/generated as part of COG-UK. COG-UK is supported by funding from the Medical Research Council (MRC) part of UK Research & Innovation (UKRI), the National Institute of Health Research (NIHR) and Genome Research Limited, operating as the Wellcome Sanger Institute. T.R.C. also acknowledges funding as part of the BBSRC Institute Strategic Programme Microbes in the Food Chain (BB/R012504/1) and its constituent projects (BBS/E/F/000PR10348 and BBS/E/F/000PR10352). J. Southgate was supported by the BBSRC -funded South West Biosciences Doctoral Training Partnership (training grant reference BB/M009122/1). AB is supported by a UKRI CDT as part of the AIMLAC CDT (grant code EP/S023992/1, project reference 2414958).

## Supplementary figure

**Figure S1.**
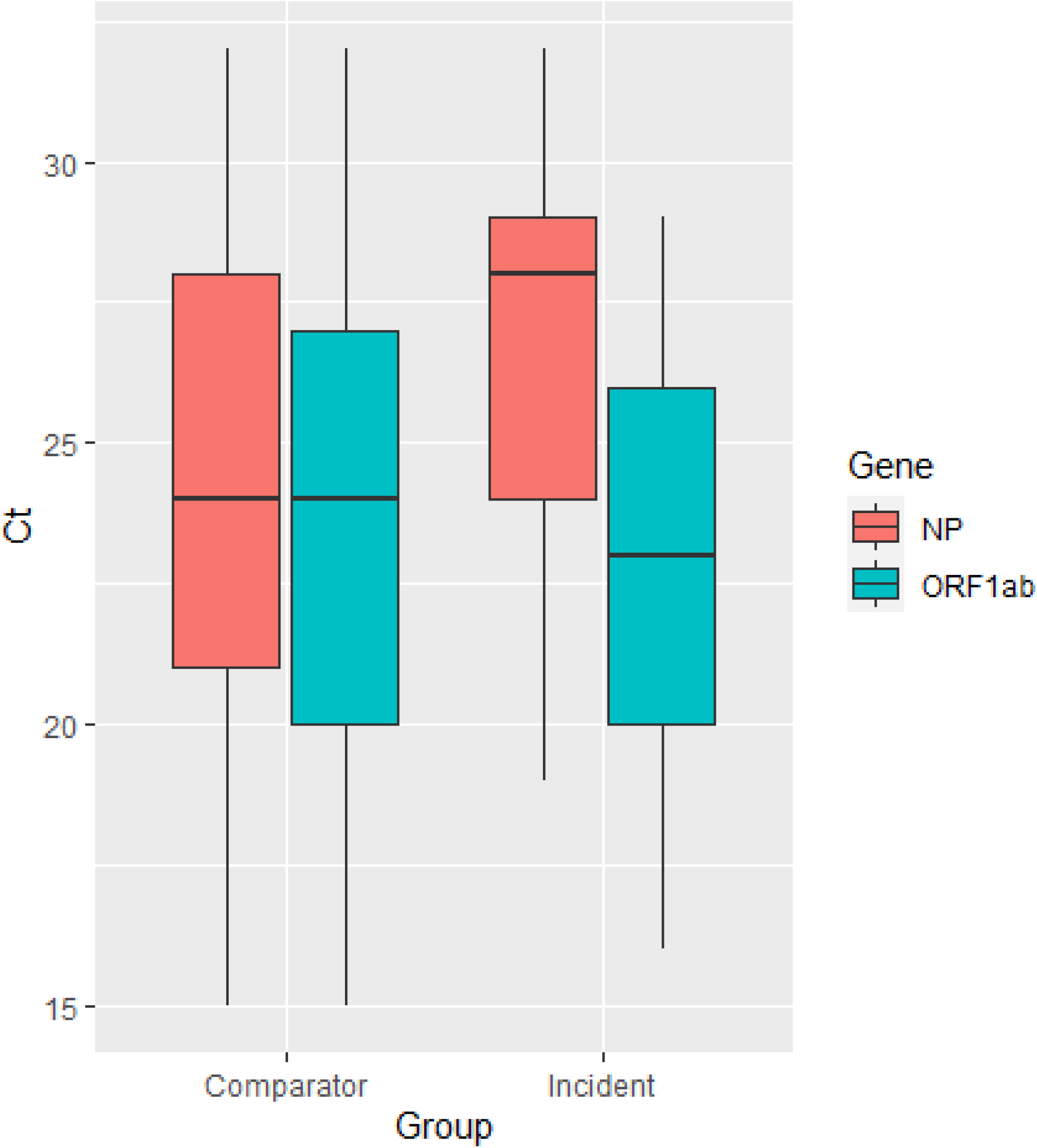
showing the distribution of Ct values for 51 samples from the incident (all of which possessed the SNPs G28881A, G28882A, G28883C and C28890T) and the distribution of Ct values from 120 positive cases, sampled over the same time period that possessed the SNPs G28881A, G28882A, G28883C but not C28890T. Data used to produce this graph is provided in Table ST2.

