## Supplementary material for "Localised community circulation of SARS-CoV-2 viruses with an increased accumulation of single nucleotide polymorphisms that adversely affect the sensitivity of real-time reverse transcription assays targeting Nucleocapsid protein": COG-UK Authorship List

**Funding acquisition, Leadership and supervision, Metadata curation, Project administration, Samples and logistics, Sequencing and analysis, Software and analysis tools, and Visualisation:** Samuel C Robson ^13^.

**Funding acquisition, Leadership and supervision, Metadata curation, Project administration, Samples and logistics, Sequencing and analysis, and Software and analysis tools:**

Nicholas J Loman ^41^ and Thomas R Connor ^10, 69^.

**Leadership and supervision, Metadata curation, Project administration, Samples and logistics, Sequencing and analysis, Software and analysis tools, and Visualisation:**

Tanya Golubchik ^5^.

**Funding acquisition, Metadata curation, Samples and logistics, Sequencing and analysis, Software and analysis tools, and Visualisation:**

Rocio T Martinez Nunez ^42^.

**Funding acquisition, Leadership and supervision, Metadata curation, Project administration, and Samples and logistics:**

Catherine Ludden ^88^.

**Funding acquisition, Leadership and supervision, Metadata curation, Samples and logistics, and Sequencing and analysis:**

Sally Corden ^69^.

**Funding acquisition, Leadership and supervision, Project administration, Samples and logistics, and Sequencing and analysis:**

Ian Johnston ^99^ and David Bonsall ^5^.

**Funding acquisition, Leadership and supervision, Sequencing and analysis, Software and analysis tools, and Visualisation:**

Colin P Smith ^87^ and Ali R Awan ^28^.

**Funding acquisition, Samples and logistics, Sequencing and analysis, Software and analysis tools, and Visualisation:**

Giselda Bucca ^87^.

**Leadership and supervision, Metadata curation, Project administration, Samples and logistics, and Sequencing and analysis:**

M. Estee Torok ^22, 101^.

**Leadership and supervision, Metadata curation, Project administration, Samples and logistics, and Visualisation:**

Kordo Saeed ^81, 110^ and Jacqui A Prieto ^83, 109^.

**Leadership and supervision, Metadata curation, Project administration, Sequencing and analysis, and Software and analysis tools:**

David K Jackson ^99^.

**Metadata curation, Project administration, Samples and logistics, Sequencing and analysis, and Software and analysis tools:**

William L Hamilton ^22^.

**Metadata curation, Project administration, Samples and logistics, Sequencing and analysis, and Visualisation:**

Luke B Snell ^11^.

**Funding acquisition, Leadership and supervision, Metadata curation, and Samples and logistics:**

Catherine Moore ^69^.

**Funding acquisition, Leadership and supervision, Project administration, and Samples and logistics:**

Ewan M Harrison ^99, 88^.

**Leadership and supervision, Metadata curation, Project administration, and Samples and logistics:**

Sonia Goncalves ^99^.

**Leadership and supervision, Metadata curation, Samples and logistics, and Sequencing and analysis:**

Ian G Goodfellow ^24^, Derek J Fairley ^3, 72^, Matthew W Loose ^18^ and Joanne Watkins ^69^.

**Leadership and supervision, Metadata curation, Samples and logistics, and Software and analysis tools:**

Rich Livett ^99^.

**Leadership and supervision, Metadata curation, Samples and logistics, and Visualisation:**

Samuel Moses ^25, 106^.

**Leadership and supervision, Metadata curation, Sequencing and analysis, and Software and analysis tools:**

Roberto Amato ^99^, Sam Nicholls ^41^ and Matthew Bull ^69^.

**Leadership and supervision, Project administration, Samples and logistics, and Sequencing and analysis:**

Darren L Smith ^37, 58, 105^.

**Leadership and supervision, Sequencing and analysis, Software and analysis tools, and Visualisation:**

Jeff Barrett ^99^, David M Aanensen ^14, 114^.

**Metadata curation, Project administration, Samples and logistics, and Sequencing and analysis:**

Martin D Curran ^65^, Surendra Parmar ^65^, Dinesh Aggarwal ^95, 99, 64^ and James G Shepherd ^48^.

**Metadata curation, Project administration, Sequencing and analysis, and Software and analysis tools:**

Matthew D Parker ^93^.

**Metadata curation, Samples and logistics, Sequencing and analysis, and Visualisation:**

Sharon Glaysher ^61^.

**Metadata curation, Sequencing and analysis, Software and analysis tools, and Visualisation:**

Matthew Bashton ^37, 58^, Anthony P Underwood ^14, 114^, Nicole Pacchiarini ^69^ and Katie F Loveson ^77^.

**Project administration, Sequencing and analysis, Software and analysis tools, and Visualisation:**

Alessandro M Carabelli ^88^.

**Funding acquisition, Leadership and supervision, and Metadata curation:**

Kate E Templeton ^53, 90^.

**Funding acquisition, Leadership and supervision, and Project administration:**

Cordelia F Langford ^99^, John Sillitoe ^99^, Thushan I de Silva ^93^ and Dennis Wang ^93^.

**Funding acquisition, Leadership and supervision, and Sequencing and analysis:**

Dominic Kwiatkowski ^99, 107^, **And**rew Rambaut ^90^, Justin O’Grady ^70, 89^ and Simon Cottrell ^69^.

**Leadership and supervision, Metadata curation, and Sequencing and analysis:**

Matthew T.G. Holden ^68^ and Emma C Thomson ^48^.

**Leadership and supervision, Project administration, and Samples and logistics:**

Husam Osman ^64, 36^, Monique Andersson ^59^, Anoop J Chauhan ^61^ and Mohammed O Hassan-Ibrahim ^6^.

**Leadership and supervision, Project administration, and Sequencing and analysis:**

Mara Lawniczak ^99^.

**Leadership and supervision, Samples and logistics, and Sequencing and analysis:**

Ravi Kumar Gupta ^88, 113^, Alex Alderton ^99^, Meera Chand ^66^, Chrystala Constantinidou ^94^, Meera Unnikrishnan ^94^, Alistair C Darby ^92^, Julian A Hiscox ^92^ and Steve Paterson ^92^.

**Leadership and supervision, Sequencing and analysis, and Software and analysis tools:**

Inigo Martincorena ^99^, David L Robertson ^48^, Erik M Volz ^39^, Andrew J Page ^70^ and Oliver G Pybus ^23^.

**Leadership and supervision, Sequencing and analysis, and Visualisation:**

Andrew R Bassett ^99^.

**Metadata curation, Project administration, and Samples and logistics:**

Cristina V Ariani ^99^, Michael H Spencer Chapman ^99, 88^, Kathy K Li ^48^, Rajiv N Shah ^48^, Natasha G Jesudason ^48^ and Yusri Taha ^50^.

**Metadata curation, Project administration, and Sequencing and analysis:**

Martin P McHugh ^53^ and Rebecca Dewar ^53^.

**Metadata curation, Samples and logistics, and Sequencing and analysis:**

Aminu S Jahun ^24^, Claire McMurray ^41^, Sarojini Pandey ^84^, James P McKenna ^3^, Andrew Nelson ^58, 105^,

Gregory R Young ^37, 58^, Clare M McCann ^58, 105^ and Scott Elliott ^61^.

**Metadata curation, Samples and logistics, and Visualisation:**

Hannah Lowe ^25^.

**Metadata curation, Sequencing and analysis, and Software and analysis tools:**

Ben Temperton ^91^, Sunando Roy ^82^, Anna Price ^10^, Sara Rey ^69^ and Matthew Wyles ^93^.

**Metadata curation, Sequencing and analysis, and Visualisation:**

Stefan Rooke ^90^ and Sharif Shaaban ^68^.

**Project administration, Samples and logistics, Sequencing and analysis:**

Mariateresa de Cesare ^98^.

**Project administration, Samples and logistics, and Software and analysis tools:**

Laura Letchford ^99^.

**Project administration, Samples and logistics, and Visualisation:**

Siona Silveira ^81^, Emanuela Pelosi ^81^ and Eleri Wilson-Davies ^81^.

**Samples and logistics, Sequencing and analysis, and Software and analysis tools:**

Myra Hosmillo ^24^.

**Sequencing and analysis, Software and analysis tools, and Visualisation:**

Áine O’Toole ^90^, Andrew R Hesketh ^87^, Richard Stark ^94^, Louis du Plessis ^23^, Chris Ruis ^88^, Helen Adams ^4^ and Yann Bourgeois ^76^.

**Funding acquisition, and Leadership and supervision:**

Stephen L Michell ^91^, Dimitris Grammatopoulos^84, 112^, Jonathan Edgeworth ^12^, Judith Breuer ^30, 82^, John A

Todd ^98^ and Christophe Fraser ^5^.

**Funding acquisition, and Project administration:**

David Buck ^98^ and Michaela John ^9^.

**Leadership and supervision, and Metadata curation:**

Gemma L Kay ^70^.

**Leadership and supervision, and Project administration:**

Steve Palmer ^99^, Sharon J Peacock ^88, 64^ and David Heyburn ^69^.

**Leadership and supervision, and Samples and logistics:**

Danni Weldon ^99^, Esther Robinson ^64, 36^, Alan McNally ^41, 86^, Peter Muir ^64^, Ian B Vipond ^64^, John BoYes ^29^, Venkat Sivaprakasam ^46^, Tranprit Salluja ^75^, Samir Dervisevic ^54^ and Emma J Meader ^54^.

**Leadership and supervision, and Sequencing and analysis:**

Naomi R Park ^99^, Karen Oliver ^99^, Aaron R Jeffries ^91^, Sascha Ott ^94^, Ana da Silva Filipe ^48^, David A Simpson ^72^ and Chris Williams ^69^.

**Leadership and supervision, and Visualisation:**

Jane AH Masoli ^73, 91^.

**Metadata curation, and Samples and logistics:**

Bridget A Knight ^73, 91^, Christopher R Jones ^73, 91^, Cherian Koshy ^1^, Amy Ash ^1^, Anna Casey ^71^, Andrew

Bosworth ^64, 36^, Liz Ratcliffe ^71^, Li Xu-McCrae ^36^, Hannah M Pymont ^64^, Stephanie Hutchings ^64^, Lisa Berry^84^, Katie Jones ^84^, Fenella Halstead ^46^, Thomas Davis ^21^, Christopher Holmes ^16^, Miren Iturriza-Gomara ^92^, Anita O Lucaci ^92^, Paul Anthony Randell ^38, 104^, Alison Cox ^38, 104^, Pinglawathee Madona ^38, 104^, Kathryn Ann Harris ^30^, Julianne Rose Brown ^30^, Tabitha W Mahungu ^74^, Dianne Irish-Tavares ^74^, Tanzina Haque ^74^, Jennifer Hart ^74^, Eric Witele ^74^, Melisa Louise Fenton ^75^, Steven Liggett ^79^, Clive Graham ^56^, Emma Swindells ^57^, Jennifer Collins ^50^, Gary Eltringham ^50^, Sharon Campbell ^17^, Patrick C McClure ^97^, Gemma Clark ^15^, Tim J Sloan ^60^, Carl Jones ^15^ and Jessica Lynch ^2, 111^.

**Metadata curation, and Sequencing and analysis:**

Ben Warne ^8^, Steven Leonard ^99^, Jillian Durham ^99^, Thomas Williams ^90^, Sam T Haldenby ^92^, Nathaniel Storey ^30^, Nabil-Fareed Alikhan ^70^, Nadine Holmes ^18^, Christopher Moore ^18^, Matthew Carlile ^18^, Malorie Perry ^69^, Noel Craine ^69^, Ronan A Lyons ^80^, Angela H Beckett ^13^, Salman Goudarzi ^77^, Christopher Fearn ^77^, Kate Cook ^77^, Hannah Dent ^77^ and Hannah Paul ^77^.

**Metadata curation, and Software and analysis tools:**

Robert Davies ^99^.

**Project administration, and Samples and logistics:**

Beth Blane ^88^, Sophia T Girgis ^88^, Mathew A Beale ^99^, Katherine L Bellis ^99, 88^, Matthew J Dorman ^99^, Eleanor Drury ^99^, Leanne Kane ^99^, Sally Kay ^99^, Samantha McGuigan ^99^, Rachel Nelson ^99^, Liam Prestwood ^99^, Shavanthi Rajatileka ^99^, Rahul Batra ^12^, Rachel J Williams ^82^, Mark Kristiansen ^82^, Angie Green ^98^, Anita Justice ^59^, Adhyana I.K Mahanama ^81, 102^ and Buddhini Samaraweera ^81, 102^.

**Project administration, and Sequencing and analysis:**

Nazreen F Hadjirin ^88^ and Joshua Quick ^41^.

**Project administration, and Software and analysis tools:**

Radoslaw Poplawski ^41^.

**Samples and logistics, and Sequencing and analysis:**

Leanne M Kermack ^88^, Nicola Reynolds ^7^, Grant Hall ^24^, Yasmin Chaudhry ^24^, Malte L Pinckert ^24^, Iliana Georgana ^24^, Robin J Moll ^99^, Alicia Thornton ^66^, Richard Myers ^66^, Joanne Stockton ^41^, Charlotte A Williams ^82^, Wen C Yew ^58^, Alexander J Trotter ^70^, Amy Trebes ^98^, George MacIntyre-Cockett ^98^, Alec Birchley ^69^, Alexander Adams ^69^, Amy Plimmer ^69^, Bree Gatica-Wilcox ^69^, Caoimhe McKerr ^69^, Ember Hilvers ^69^, Hannah Jones ^69^, Hibo Asad ^69^, Jason Coombes ^69^, Johnathan M Evans ^69^, Laia Fina ^69^, Lauren Gilbert ^69^, Lee Graham ^69^, Michelle Cronin ^69^, Sara Kumziene-SummerhaYes ^69^, Sarah Taylor ^69^, Sophie Jones ^69^, Danielle C Groves ^93^, Peijun Zhang ^93^, Marta Gallis ^93^ and Stavroula F Louka ^93^.

**Samples and logistics, and Software and analysis tools:**

Igor Starinskij ^48^.

**Sequencing and analysis, and Software and analysis tools:**

Chris J Illingworth ^47^, Chris Jackson ^47^, Marina Gourtovaia ^99^, Gerry Tonkin-Hill ^99^, Kevin Lewis ^99^, Jaime M Tovar-Corona ^99^, Keith James ^99^, Laura Baxter ^94^, Mohammad T. Alam ^94^, Richard J Orton ^48^, Joseph Hughes ^48^, Sreenu Vattipally ^48^, Manon Ragonnet-Cronin ^39^, Fabricia F. Nascimento ^39^, David Jorgensen ^39^, Olivia Boyd ^39^, Lily Geidelberg ^39^, Alex E Zarebski ^23^, Jayna Raghwani ^23^, Moritz UG Kraemer ^23^, Joel Southgate ^10, 69^, Benjamin B Lindsey ^93^ and Timothy M Freeman ^93^.

**Software and analysis tools, and Visualisation:**

Jon-Paul Keatley ^99^, Joshua B Singer ^48^, Leonardo de Oliveira Martins ^70^, Corin A Yeats ^14^, Khalil Abudahab ^14, 114^, Ben EW Taylor ^14, 114^ and Mirko Menegazzo ^14^.

**Leadership and supervision:**

John Danesh ^99^, Wendy Hogsden ^46^, Sahar Eldirdiri ^21^, Anita Kenyon ^21^, Jenifer Mason ^43^, Trevor I Robinson ^43^, Alison Holmes ^38, 103^, James Price ^38, 103^, John A Hartley ^82^, Tanya Curran ^3^, Alison E Mather ^70^, Giri Shankar ^69^, Rachel Jones ^69^, Robin Howe ^69^ and Sian Morgan ^9^.

**Metadata curation:**

Elizabeth Wastenge ^53^, Michael R Chapman ^34, 88, 99^, Siddharth Mookerjee ^38, 103^, Rachael Stanley ^54^, Wendy Smith ^15^, Timothy Peto ^59^, David Eyre ^59^, Derrick Crook ^59^, Gabrielle Vernet ^33^, Christine Kitchen ^10^, Huw Gulliver ^10^, Ian Merrick ^10^, Martyn Guest ^10^, Robert Munn ^10^, Declan T Bradley ^63, 72^ and Tim Wyatt ^63^.

**Project administration:**

Charlotte Beaver ^99^, Luke Foulser ^99^, Sophie Palmer ^88^, Carol M Churcher ^88^, Ellena Brooks ^88^, Kim S Smith ^88^, Katerina Galai ^88^, Georgina M McManus ^88^, Frances Bolt ^38, 103^, Francesc Coll ^19^, Lizzie Meadows ^70^, Stephen W Attwood ^23^, Alisha Davies ^69^, Elen De Lacy ^69^, Fatima Downing ^69^, Sue Edwards ^69^, Garry P Scarlett ^76^, Sarah Jeremiah ^83^ and Nikki Smith ^93^.

**Samples and logistics:**

Danielle Leek ^88^, Sushmita Sridhar ^88, 99^, Sally Forrest ^88^, Claire Cormie ^88^, Harmeet K Gill ^88^, Joana Dias ^88^, Ellen E Higginson ^88^, Mailis Maes ^88^, Jamie Young ^88^, Michelle Wantoch ^7^, Sanger Covid Team ([www.sanger.ac.uk/covid-team](http://www.sanger.ac.uk/covid-team)) ^99^, Dorota Jamrozy ^99^, Stephanie Lo ^99^, Minal Patel ^99^, Verity Hill ^90^, Claire M Bewshea ^91^, Sian Ellard ^73, 91^, Cressida Auckland ^73^, Ian Harrison ^66^, Chloe Bishop ^66^, Vicki Chalker ^66^, Alex Richter ^85^, Andrew Beggs ^85^, Angus Best ^86^, Benita Percival ^86^, Jeremy Mirza ^86^, Oliver Megram ^86^, Megan Mayhew ^86^, Liam Crawford ^86^, Fiona Ashcroft ^86^, Emma Moles-Garcia ^86^, Nicola Cumley ^86^, Richard Hopes ^64^, Patawee Asamaphan ^48^, Marc O Niebel ^48^, Rory N Gunson ^100^, Amanda Bradley ^52^, Alasdair Maclean ^52^, Guy Mollett ^52^, Rachel Blacow ^52^, Paul Bird ^16^, Thomas Helmer ^16^, Karlie Fallon ^16^, Julian Tang ^16^, Antony D Hale ^49^, Louissa R Macfarlane-Smith ^49^, Katherine L Harper ^49^, Holli Carden ^49^, Nicholas W Machin ^45, 64^, Kathryn A Jackson ^92^, Shazaad S Y Ahmad ^45, 64^, Ryan P George ^45^, Lance Turtle ^92^, Elaine O’Toole ^43^, Joanne Watts ^43^, Cassie Breen ^43^, Angela Cowell ^43^, Adela Alcolea-Medina ^32, 96^, Themoula Charalampous ^12, 42^, Amita Patel ^11^, Lisa J Levett ^35^, Judith Heaney ^35^, Aileen Rowan ^39^, Graham P Taylor ^39^, Divya Shah ^30^, Laura Atkinson ^30^, Jack CD Lee ^30^, Adam P Westhorpe ^82^, Riaz Jannoo ^82^, Helen L Lowe ^82^, Angeliki Karamani ^82^, Leah Ensell ^82^, Wendy Chatterton ^35^, Monika Pusok ^35^, Ashok Dadrah ^75^, Amanda Symmonds ^75^, Graciela Sluga ^44^, Zoltan Molnar ^72^, Paul Baker ^79^, Stephen Bonner ^79^, Sarah Essex ^79^, Edward Barton ^56^, Debra Padgett ^56^, Garren Scott ^56^, Jane Greenaway ^57^, Brendan AI Payne ^50^, Shirelle Burton-Fanning ^50^, Sheila Waugh ^50^, Veena Raviprakash ^17^, Nicola Sheriff ^17^, Victoria Blakey ^17^, Lesley-Anne Williams ^17^, Jonathan Moore ^27^, Susanne Stonehouse ^27^, Louise Smith ^55^, Rose K Davidson ^89^, Luke Bedford ^26^, Lindsay Coupland ^54^, Victoria Wright ^18^, Joseph G Chappell ^97^, Theocharis Tsoleridis ^97^, Jonathan Ball ^97^, Manjinder Khakh ^15^, Vicki M Fleming ^15^, Michelle M Lister ^15^, Hannah C Howson-Wells ^15^, Louise Berry ^15^, Tim Boswell ^15^, Amelia Joseph ^15^, Iona Willingham ^15^, Nichola Duckworth ^60^, Sarah Walsh ^60^, Emma Wise ^2, 111^, Nathan Moore ^2, 111^, Matilde Mori ^2, 108, 111^, Nick Cortes ^2, 111^, Stephen Kidd ^2,111^, Rebecca Williams ^33^, Laura Gifford ^69^, Kelly Bicknell ^61^, Sarah Wyllie ^61^, Allyson Lloyd ^61^, Robert Impey ^61^, Cassandra S Malone ^6^, Benjamin J Cogger ^6^, Nick Levene ^62^, Lynn Monaghan ^62^, Alexander J Keeley ^93^, David G Partridge ^78, 93^, Mohammad Raza ^78, 93^, Cariad Evans ^78, 93^ and Kate Johnson ^78, 93^.

**Sequencing and analysis:**

Emma Betteridge ^99^, Ben W Farr ^99^, Scott Goodwin ^99^, Michael A Quail ^99^, Carol Scott ^99^, Lesley Shirley ^99^, Scott AJ Thurston ^99^, Diana Rajan ^99^, Iraad F Bronner ^99^, Louise Aigrain ^99^, Nicholas M Redshaw ^99^, Stefanie V Lensing ^99^, Shane McCarthy ^99^, Alex Makunin ^99^, Carlos E Balcazar ^90^, Michael D Gallagher ^90^, Kathleen A Williamson ^90^, Thomas D Stanton ^90^, Michelle L Michelsen ^91^, Joanna Warwick-Dugdale ^91^, Robin Manley ^91^, Audrey Farbos ^91^, James W Harrison ^91^, Christine M Sambles ^91^, David J Studholme ^91^, Angie Lackenby ^66^, Tamyo Mbisa ^66^, Steven Platt ^66^, Shahjahan Miah ^66^, David Bibby ^66^, Carmen Manso ^66^, Jonathan Hubb ^66^, Gavin Dabrera ^66^, Mary Ramsay ^66^, Daniel Bradshaw ^66^, Ulf Schaefer ^66^, Natalie Groves ^66^, Eileen Gallagher ^66^, David Lee ^66^, David Williams ^66^, Nicholas Ellaby ^66^, Hassan Hartman ^66^, Nikos Manesis ^66^, Vineet Patel ^66^, Juan Ledesma ^67^, Katherine A Twohig ^67^, Elias Allara ^64, 88^, Clare Pearson ^64, 88^, Jeffrey K. J. Cheng ^94^, Hannah E. Bridgewater ^94^, Lucy R. Frost ^94^, Grace Taylor-Joyce ^94^, Paul E Brown ^94^, Lily Tong ^48^, Alice Broos ^48^, Daniel Mair ^48^, Jenna Nichols ^48^, Stephen N Carmichael ^48^, Katherine L Smollett ^40^, Kyriaki Nomikou ^48^, Elihu Aranday-Cortes ^48^, Natasha Johnson ^48^, Seema Nickbakhsh ^48, 68^, Edith E Vamos ^92^, Margaret Hughes ^92^, Lucille Rainbow ^92^, Richard Eccles ^92^, Charlotte Nelson ^92^, Mark Whitehead ^92^, Richard Gregory ^92^, Matthew Gemmell ^92^, Claudia Wierzbicki ^92^, Hermione J Webster ^92^, Chloe L Fisher ^28^, Adrian W Signell ^20^, Gilberto Betancor ^20^, Harry D Wilson ^20^, Gaia Nebbia ^12^, Flavia Flaviani ^31^, Alberto C Cerda ^96^, Tammy V Merrill ^96^, Rebekah E Wilson ^96^, Marius Cotic ^82^, Nadua Bayzid ^82^, Thomas Thompson ^72^, Erwan Acheson ^72^, Steven Rushton ^51^, Sarah O’Brien ^51^, David J Baker ^70^, Steven Rudder ^70^, Alp Aydin ^70^, Fei Sang ^18^, Johnny Debebe ^18^, Sarah Francois ^23^, Tetyana I Vasylyeva ^23^, Marina Escalera Zamudio ^23^, Bernardo Gutierrez ^23^, Angela Marchbank ^10^, Joshua Maksimovic ^9^, Karla Spellman ^9^, Kathryn McCluggage ^9^, Mari Morgan ^69^, Robert Beer ^9^, Safiah Afifi ^9^, Trudy Workman ^10^, William Fuller^10^, Catherine Bresner ^10^, Adrienn Angyal ^93^, Luke R Green ^93^, Paul J Parsons ^93^, Rachel M Tucker ^93^, Rebecca Brown ^93^ and Max Whiteley ^93^.

**Software and analysis tools:**

James Bonfield ^99^, Christoph Puethe ^99^, Andrew Whitwham ^99^, Jennifier Liddle ^99^, Will Rowe ^41^, Igor Siveroni ^39^, Thanh Le-Viet ^70^ and Amy Gaskin ^69^.

**Visualisation:**

Rob Johnson ^39^.

**1** Barking, Havering and Redbridge University Hospitals NHS Trust, **2** Basingstoke Hospital, **3** Belfast Health & Social Care Trust, **4** Betsi Cadwaladr University Health Board, **5** Big Data Institute, Nuffield Department of Medicine, University of Oxford, **6** Brighton and Sussex University Hospitals NHS Trust, **7** Cambridge Stem Cell Institute, University of Cambridge, **8** Cambridge University Hospitals NHS Foundation Trust, **9** Cardiff and Vale University Health Board, **10** Cardiff University, **11** Centre for Clinical Infection & Diagnostics Research, St. Thomas’ Hospital and Kings College London, **12** Centre for Clinical Infection and Diagnostics Research, Department of Infectious Diseases, Guy’s and St Thomas’ NHS Foundation Trust, **13** Centre for Enzyme Innovation, University of Portsmouth (PORT), **14** Centre for Genomic Pathogen Surveillance, University of Oxford, **15** Clinical Microbiology Department, Queens Medical Centre, **16** Clinical Microbiology, University Hospitals of Leicester NHS Trust, **17** County Durham and Darlington NHS Foundation Trust, **18** Deep Seq, School of Life Sciences, Queens Medical Centre, University of Nottingham, **19** Department of Infection Biology, Faculty of Infectious & Tropical Diseases, London School of Hygiene & Tropical Medicine, **20** Department of Infectious Diseases, King’s College London, **21** Department of Microbiology, Kettering General Hospital, **22** Departments of Infectious Diseases and Microbiology, Cambridge University Hospitals NHS Foundation Trust; Cambridge, UK, **23** Department of Zoology, University of Oxford, **24** Division of Virology, Department of Pathology, University of Cambridge, **25** East Kent Hospitals University NHS Foundation Trust, **26** East Suffolk and North Essex NHS Foundation Trust, **27** Gateshead Health NHS Foundation Trust, **28** Genomics Innovation Unit, Guy’s and St. Thomas’ NHS Foundation Trust, **29** Gloucestershire Hospitals NHS Foundation Trust, **30** Great Ormond Street Hospital for Children NHS Foundation Trust, **31** Guy’s and St. Thomas’ BRC, **32** Guy’s and St. Thomas’ Hospitals, **33** Hampshire Hospitals NHS Foundation Trust, **34** Health Data Research UK Cambridge, **35** Health Services Laboratories, **36** Heartlands Hospital, Birmingham, **37** Hub for Biotechnology in the Built Environment, Northumbria University, **38** Imperial College Hospitals NHS Trust, **39** Imperial College London, **40** Institute of Biodiversity, Animal Health & Comparative Medicine, **41** Institute of Microbiology and Infection, University of Birmingham, **42** King’s College London, **43** Liverpool Clinical Laboratories, **44** Maidstone and Tunbridge Wells NHS Trust, **45** Manchester University NHS Foundation Trust, **46** Microbiology Department, Wye Valley NHS Trust, Hereford, **47** MRC Biostatistics Unit, University of Cambridge, **48** MRC-University of Glasgow Centre for Virus Research, **49** National Infection Service, PHE and Leeds Teaching Hospitals Trust, **50** Newcastle Hospitals NHS Foundation Trust, **51** Newcastle University, **52** NHS Greater Glasgow and Clyde, **53** NHS Lothian, **54** Norfolk and Norwich University Hospital, **55** Norfolk County Council, **56** North Cumbria Integrated Care NHS Foundation Trust, **57** North Tees and Hartlepool NHS Foundation Trust, **58** Northumbria University, **59** Oxford University Hospitals NHS Foundation Trust, **60** PathLinks, Northern Lincolnshire & Goole NHS Foundation Trust, **61** Portsmouth Hospitals University NHS Trust, **62** Princess Alexandra Hospital Microbiology Dept., **63** Public Health Agency, **64** Public Health England, **65** Public Health England, Clinical Microbiology and Public Health Laboratory, Cambridge, UK, **66** Public Health England, Colindale, **67** Public Health England, Colindale, **68** Public Health Scotland, **69** Public Health Wales NHS Trust, **70** Quadram Institute Bioscience, **71** Queen Elizabeth Hospital, **72** Queen’s University Belfast, **73** Royal Devon and Exeter NHS Foundation Trust, **74** Royal Free NHS Trust, **75** Sandwell and West Birmingham NHS Trust, **76** School of Biological Sciences, University of Portsmouth (PORT), **77** School of Pharmacy and Biomedical Sciences, University of Portsmouth (PORT), **78** Sheffield Teaching Hospitals, **79** South Tees Hospitals NHS Foundation Trust, **80** Swansea University, **81** University Hospitals Southampton NHS Foundation Trust, **82** University College London, **83** University Hospital Southampton NHS Foundation Trust, **84** University Hospitals Coventry and Warwickshire, **85** University of Birmingham, **86** University of Birmingham Turnkey Laboratory, **87** University of Brighton, **88** University of Cambridge, **89** University of East Anglia, **90** University of Edinburgh, **91** University of Exeter, **92** University of Liverpool, **93** University of Sheffield, **94** University of Warwick, **95** University of Cambridge, **96** Viapath, Guy’s and St Thomas’ NHS Foundation Trust, and King’s College Hospital NHS Foundation Trust, **97** Virology, School of Life Sciences, Queens Medical Centre, University of Nottingham, **98** Wellcome Centre for Human Genetics, Nuffield Department of Medicine, University of Oxford, **99** Wellcome Sanger Institute, **100** West of Scotland Specialist Virology Centre, NHS Greater Glasgow and Clyde, **101** Department of Medicine, University of Cambridge, **102** Ministry of Health, Sri Lanka, **103** NIHR Health Protection Research Unit in HCAI and AMR, Imperial College London, **104** North West London Pathology, **105** NU-OMICS, Northumbria University, **106** University of Kent, **107** University of Oxford, **108** University of Southampton, **109** University of Southampton School of Health Sciences, **110** University of Southampton School of Medicine, **111** University of Surrey, **112** Warwick Medical School and Institute of Precision Diagnostics, Pathology, UHCW NHS Trust, **113** Wellcome Africa Health Research Institute Durban and **114** Wellcome Genome Campus.
